# Attenuated alpha state transitions and disrupted repetition-dependent temporal adaptation in mild cognitive impairment

**DOI:** 10.64898/2026.10.10.26365211

**Authors:** Xuan Wang, Ying Zhu, Jian Zhang, Zheng Zheng, Ming Yu

## Abstract

Mild cognitive impairment (MCI) may involve impaired dynamic regulation of brain states that is not captured by conventional resting-state electroencephalography (EEG). We quantified occipital alpha (8–12 Hz) transitions during three repeated eye-opening/eye-closure cycles in 60 healthy controls (HC) and 77 participants with MCI. Eye-state events were refined from frontal ocular artifacts. Transition magnitude was measured by the log-transformed post-/pre-event energy ratio, and transition success by the expected direction of alpha change. For canonical transitions meeting prespecified fit-quality criteria, mono-exponential fitting yielded transition onset (t_onset_) and the post-onset time constant (τ). MCI showed weaker alpha suppression during eye opening, weaker alpha enhancement during eye closure, and lower odds of transition success. No significant group difference in τ was detected within the canonical quality-controlled subset. In contrast, t_onset_ showed significant Group × Trial interactions in both conditions: HC developed later onset during eye opening and earlier onset during eye closure across repetitions, whereas MCI showed no comparable change. These findings suggest reduced alpha state-transition capacity and altered repetition-dependent temporal organization in MCI.

## Introduction

Mild cognitive impairment (MCI) is an important clinical stage between normal cognitive ageing and dementia, with heterogeneous outcomes but an increased risk of progression to Alzheimer’s disease and other dementias. Contemporary clinical assessment integrates history, objective cognitive testing and preservation of functional independence, with imaging or molecular biomarkers used when clinically or scientifically appropriate [1,2]. Electroencephalography (EEG), with millisecond-scale temporal resolution and direct sensitivity to neural population activity, is well suited to characterize fast changes in brain state. Recent primary studies have identified abnormal resting-state oscillations and functional connectivity in MCI [3], while task-based and reduced-electrode EEG approaches have also shown discriminative signatures across the MCI/Alzheimer disease spectrum [4,5]. These observations support EEG as a complementary approach for probing functional regulation that is not captured by global cognitive scores alone.

Eye opening and eye closure evoke one of the most reproducible changes in human EEG: posterior alpha activity decreases when the eyes open and re-emerges when the eyes close [6,7]. Recent work further indicates that alpha activity tracks shifts between externally and internally oriented processing modes [8]. Reduced alpha reactivity to eye opening has been reported in MCI and Alzheimer’s disease [9], and diminished eyes-closed-to-eyes-open reactivity has also been associated with cerebral amyloid burden in non-demented older adults [10]. More recently, posterior alpha reactivity has been shown to vary with short-term progression in Alzheimer disease-related MCI [11]. Together, these findings suggest that the eye-state response contains clinically relevant information beyond static alpha power. However, most prior work has summarized power or reactivity across relatively long eye-state periods, leaving the transition trajectory itself—whether the expected response occurs reliably, how large it is, when it begins relative to the eye-state event, and how rapidly it evolves once initiated—less well characterized.

Viewing eye-state change as a perturbation of brain state offers a complementary framework for investigating dynamic neural regulation in MCI. Recent studies have reported altered EEG microstate dynamics [12] and resting-state network organization across MCI subtypes [13], as well as abnormal alpha reactivity during vigilance-state transitions in Alzheimer’s disease-related MCI [14,15]. During cognitive tasks, MCI has also been associated with altered alpha/beta network organization [16], theta/alpha functional connectivity [17], and directed frontal–posterior and interhemispheric connectivity [18]. Longitudinal studies further suggest that quantitative EEG (qEEG) slowing, holo-spectral EEG features, functional connectivity, and multifeature EEG models may be associated with subsequent progression from MCI to dementia or Alzheimer’s disease [19–22]. Recent Scientific Reports studies have additionally demonstrated altered resting-state EEG network activity in Alzheimer’s disease [23] and the feasibility of selecting informative EEG channels and features for distinguishing MCI from healthy controls [24]. Together, these findings suggest that EEG abnormalities in MCI extend beyond static signal differences to alterations in the organization and modulation of neural activity; however, how the brain executes a simple physiological state transition, and whether its temporal organization changes when the same transition is repeated, remain less well characterized. Repeated eye opening and eye closure provide a simple within-person paradigm with which to address this gap by quantifying transition magnitude, transition-success probability, fitted onset (t_onset_), and post-onset time constant (τ), and by determining whether these features change across repeated trials.

We therefore examined occipital alpha transitions during three consecutive eye-opening/eye-closure cycles in healthy controls (HC) and MCI. We asked whether MCI was associated with reduced transition magnitude or a lower probability of canonical alpha modulation; whether τ or t_onset_ differed among canonical quality-controlled transitions; whether these measures changed differently across repeated trials; and whether repetition-related changes were associated with Montreal Cognitive Assessment (MoCA) scores. We hypothesized that MCI would show impaired expression of alpha-state transitions and altered repetition-dependent temporal organization. The analysis framework is summarized in Fig. 1.

**Figure 1.**
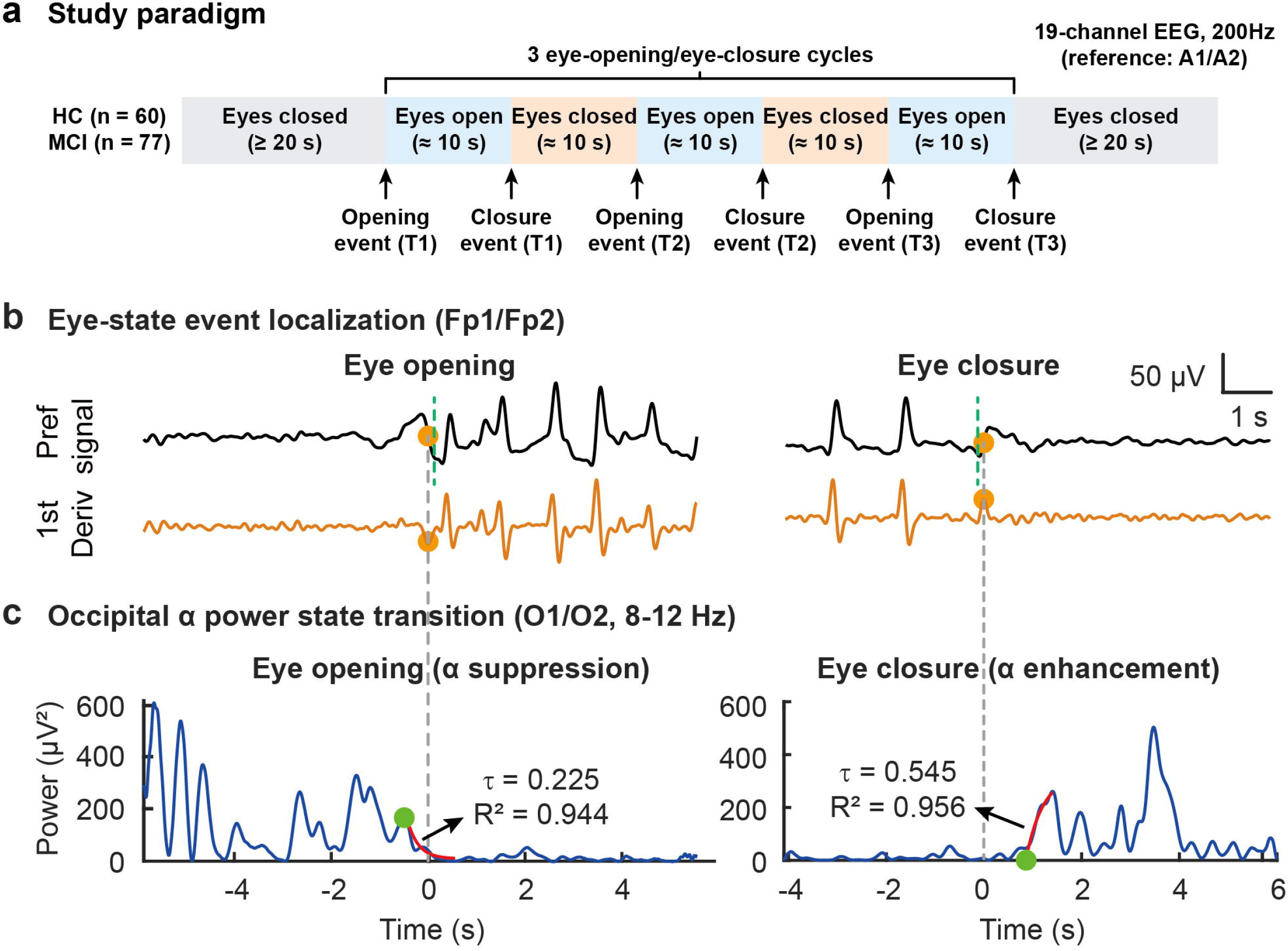
Experimental paradigm and analysis framework for eye-state alpha transitions. a, Eye-state paradigm for 60 healthy controls (HC) and 77 participants with mild cognitive impairment (MCI): 19-channel EEG at 200 Hz, an initial eyes-closed period, three approximately 10-s eye-opening/eye-closure cycles, and a final eyes-closed period. T1–T3 denote successive transitions of the same type. b, Representative event localization from the averaged Fp1/Fp2 ocular proxy (black) and its first derivative (orange). Green dashed lines mark the examiner-entered event markers; grey dashed lines mark the refined ocular events used as t=0 and continue into panel c. c, Example single-trial O1/O2 alpha-energy trajectories (8–12 Hz) for eye opening and closure. Blue traces show measured energy, red segments the mono-exponential fits, and green circles fitted t_onset_; labels show the fitted post-onset time constant (τ).

**Figure 2.**
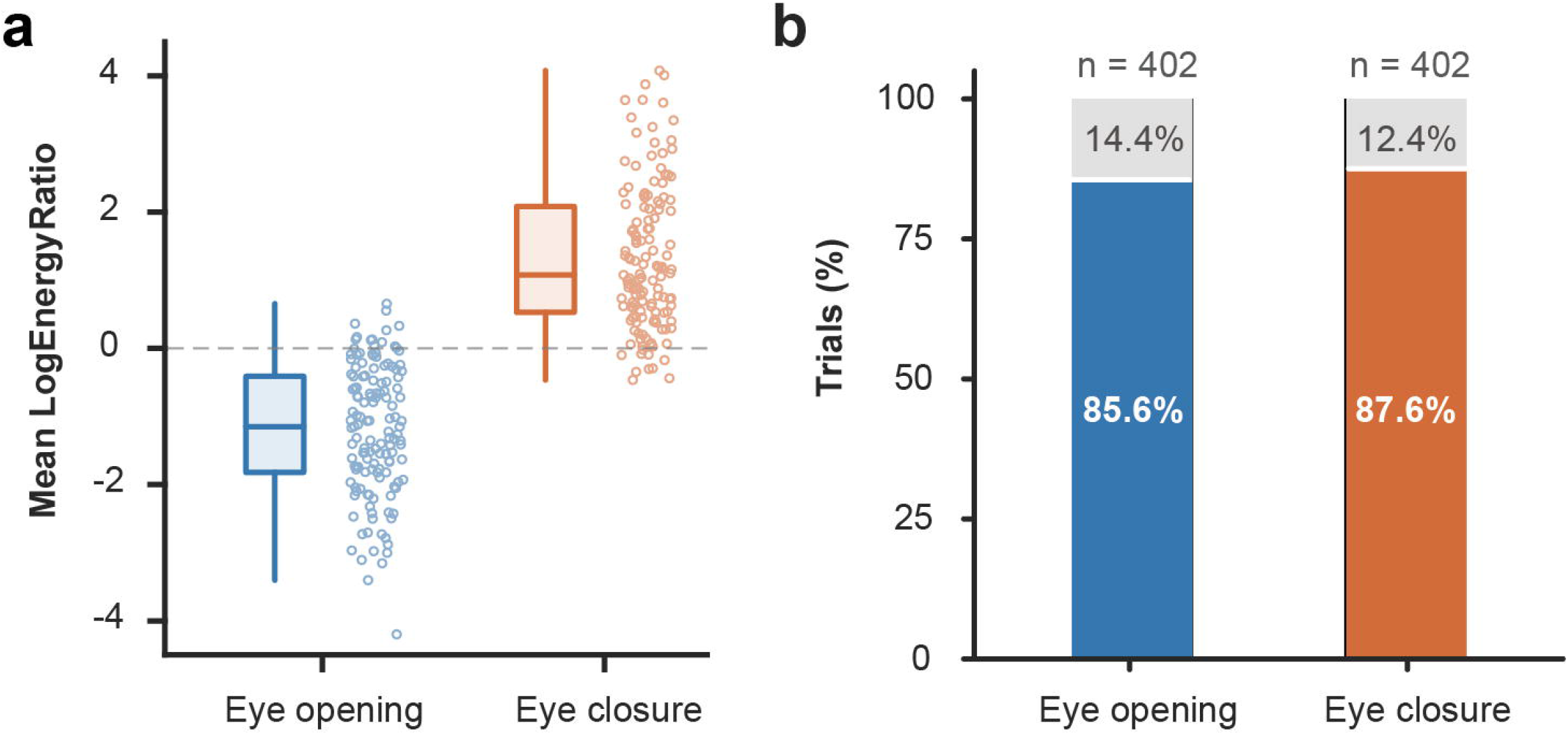
Validation of the bidirectional alpha state-transition paradigm. a, Participant-level mean LogEnergyRatio across available Trials 1–3 for eye opening and closure (HC, n=60; MCI, n=77). Each point represents one participant; boxes show the median and interquartile range, with whiskers extending to the most extreme values within 1.5×IQR. The dashed line denotes no net energy change. The expected direction was observed in 90.5% of participants for opening and 92.7% for closure (two-sided Wilcoxon signed-rank tests versus zero, both P<0.001). b, Trial-level transition success among 402 eye-opening and 402 eye-closure transitions: 85.6% for opening (EnergyRatio<1) and 87.6% for closure (EnergyRatio>1). Colored segments show successful transitions and grey segments the remainder.

**Figure 3.**
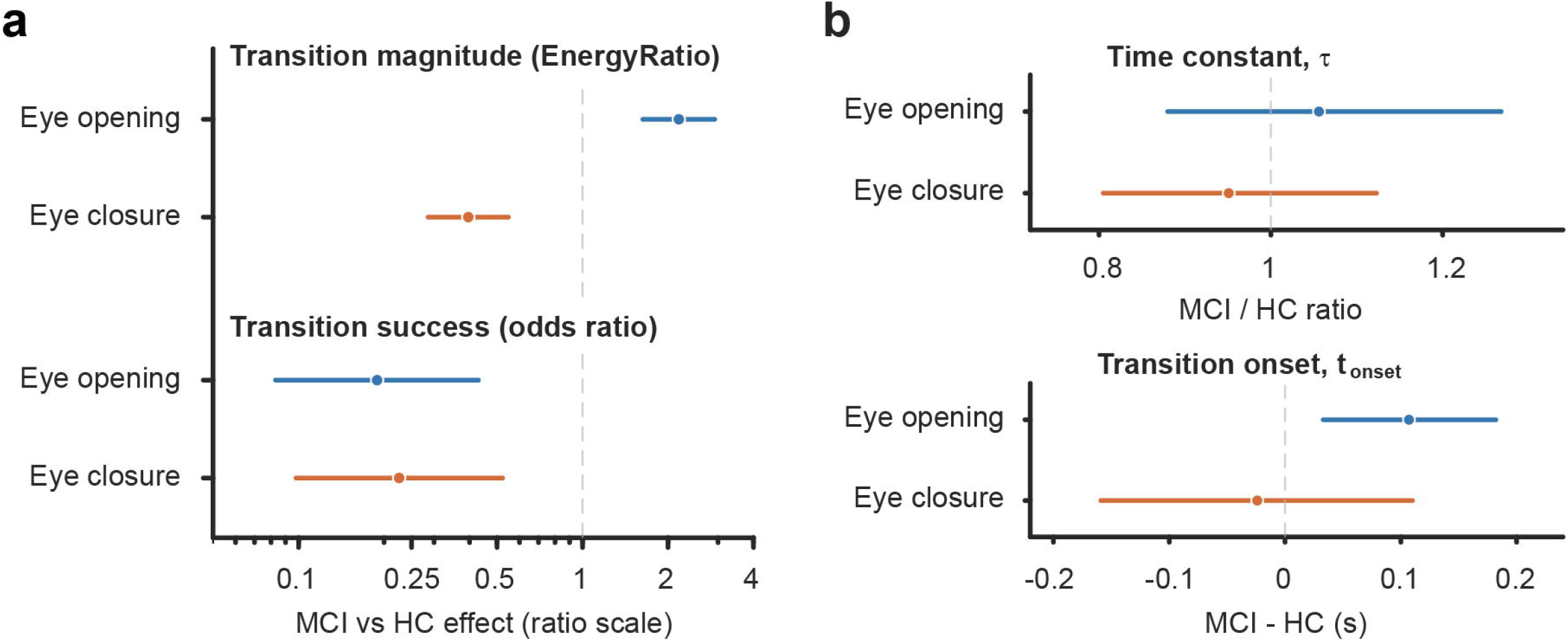
Adjusted group differences in alpha state-transition capacity and kinetics. a, Covariate-adjusted MCI-versus-HC effects on transition magnitude and transition success during eye opening (blue) and closure (orange). Magnitude is shown as the MCI/HC EnergyRatio ratio derived from LogEnergyRatio models; success is shown as an odds ratio. The null ratio is 1. b, Conditional kinetic effects among canonical quality-controlled transitions (opening, n=275; closure, n=278): the MCI/HC ratio of τ and the adjusted MCI-minus-HC difference in t_onset_. Null values are 1 and 0, respectively. Points show adjusted estimates and bars 95% confidence intervals. Models adjusted for age, sex, education and Trial with a participant-specific random intercept; Holm-adjusted P values are reported in Table 2.

**Figure 4.**
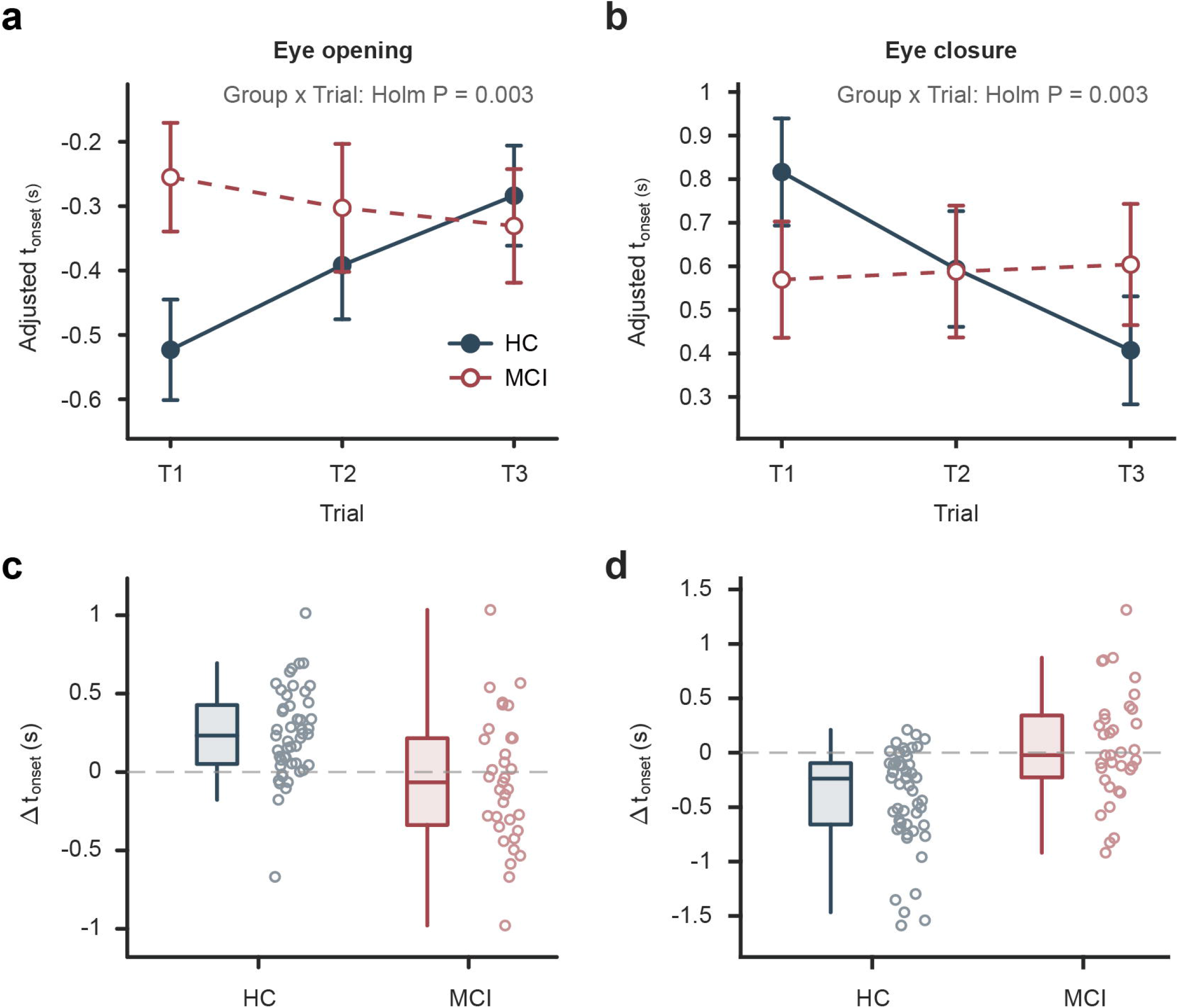
Altered repetition-dependent adaptation of alpha state-transition onset timing in MCI. **a,b**, Covariate-adjusted marginal t_onset_ estimates across Trials 1–3 for eye opening (a) and eye closure(b) in HC and MCI. Points and bars show estimates and 95% confidence intervals; Group × Trial interactions were significant in both conditions (Holm-adjusted P=0.003 each). **c,d**, Participant-level T3−T1 changes in t_onset_ for eye opening (c; HC n=49, MCI n=35) and eye closure (d; HC n=53, MCI n=35), restricted to participants with canonical quality-controlled fits at both trials. Each point represents one participant; boxes show the median and interquartile range, with whiskers extending to the most extreme values within 1.5×IQR. The dashed line denotes no change.

## Results

### Participant characteristics and paradigm validation

The study included 137 participants (60 HC and 77 MCI). Age, sex and years of education did not differ significantly between groups (Table 1), whereas MoCA and Mini-Mental State Examination (MMSE) scores were lower in MCI (both P<0.001). Clinical Dementia Rating (CDR) distributions are summarized descriptively in Table 1. Across all participants, mean LogEnergyRatio, defined as the natural logarithm of the post-/pre-event alpha-energy ratio, was negative after eye opening (-1.20±0.96) and positive after eye closure (1.31±1.07), both P<0.001. At the participant level, 90.5% showed the expected direction after eye opening and 92.7% after eye closure. At the trial level, 402 eye-opening and 402 eye-closure transitions were available; transition-success rates were 85.6% and 87.6%, respectively. Group-specific transition-success rates were 95.0% for both conditions in HC, compared with 78.0% for eye opening and 81.6% for eye closure in MCI (Supplementary Table S1).

**Table 1.**
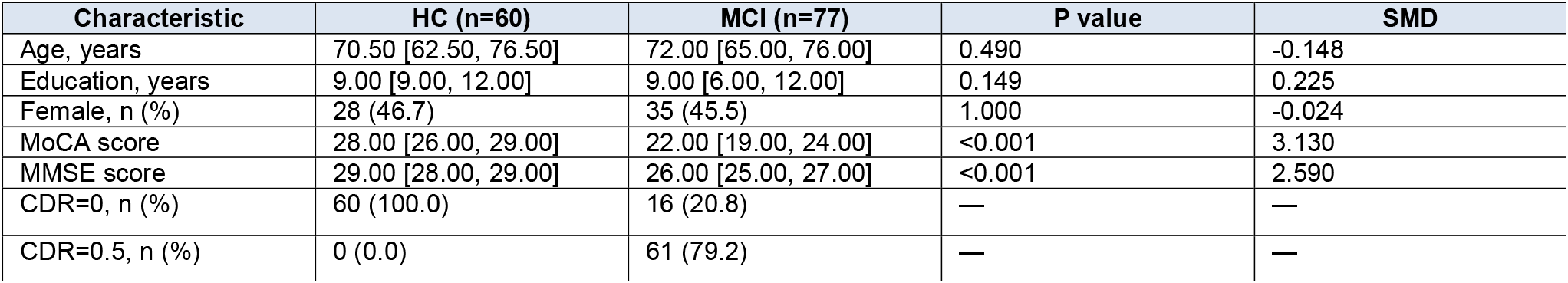
Participant demographic and cognitive characteristics. Data are median [interquartile range] or n (%). MoCA scores include the standard 1-point education adjustment for participants with ≤12 years of education. CDR is reported descriptively. HC, healthy controls; MCI, mild cognitive impairment; MoCA, Montreal Cognitive Assessment; MMSE, Mini-Mental State Examination; CDR, Clinical Dementia Rating; SMD, standardized mean difference.

### Reduced alpha state-transition capacity in MCI

In covariate-adjusted mixed-effects models, MCI showed weaker alpha modulation in both conditions. During eye opening, LogEnergyRatio was higher in MCI than HC (β=0.781, 95% confidence interval [CI] 0.489–1.072; Holm-adjusted P<0.001), corresponding to a 2.18-fold higher EnergyRatio, the untransformed post-/pre-event alpha-energy ratio (95% CI 1.63–2.92); median EnergyRatio was 0.178 in HC and 0.536 in MCI. Because the expected eye-opening response is suppression, values closer to 1 indicate weaker modulation. During eye closure, LogEnergyRatio was lower in MCI (β=-0.926, 95% CI - 1.250 to -0.601; Holm-adjusted P<0.001), corresponding to an EnergyRatio ratio of 0.396 (95% CI 0.286–0.548); median EnergyRatio was 5.88 in HC and 1.91 in MCI. Thus, MCI showed attenuated modulation in both directions; adjusted effects and descriptive EnergyRatio medians are reported in Table 2 and Supplementary Table S1C, respectively.

**Table 2.** Adjusted group differences in alpha state-transition measures and conditional kinetics. Mixed-effects models adjusted for age, sex, education and Trial, with participant-specific random intercepts. β is the adjusted MCI-versus-HC coefficient for LogEnergyRatio and t_onset_; OR is the odds ratio for TransitionSuccess; exp(β) for LogTau is reported as the MCI/HC ratio on the original τ scale. Kinetic outcomes were analyzed only among canonical quality-controlled transitions. Holm-adjusted P values follow the prespecified outcome families defined in the Methods. CI, confidence interval.

| Condition | Outcome | Analysis set | Adjusted effect estimate (95% CI) | Holm-adjusted P |
| --- | --- | --- | --- | --- |
| Open | LogEnergyRatio | 402 trials / 137 participants | $\beta=0.781$ (0.489, 1.072) | <0.001 |
| Close | LogEnergyRatio | 402 trials / 137 participants | $\beta=-0.926$ (-1.250, -0.601) | <0.001 |
| Open | TransitionSuccess | 402 trials / 137 participants | OR=0.189 (0.083, 0.430) | <0.001 |
| Close | TransitionSuccess | 402 trials / 137 participants | OR=0.226 (0.098, 0.523) | <0.001 |
| Open | LogTau | 275 trials / 115 participants | MCI/HC ratio=1.056 (0.880, 1.268) | 1.000 |
| Close | LogTau | 278 trials / 116 participants | MCI/HC ratio=0.951 (0.805, 1.123) | 1.000 |
| Open | $t_{\text{onset}}$, s | 275 trials / 115 participants | $\beta=0.107$ (0.033, 0.182) | 0.020 |
| Close | $t_{\text{onset}}$, s | 278 trials / 116 participants | $\beta=-0.024$ (-0.159, 0.110) | 1.000 |

MCI also had substantially lower odds of transition success during eye opening (adjusted odds ratio [OR]=0.189, 95% CI 0.083–0.430; Holm-adjusted P<0.001) and eye closure (OR=0.226, 95% CI 0.098–0.523; Holm-adjusted P<0.001), indicating that the group difference involved both transition magnitude and the probability of producing the expected physiological response (Table 2).

### Conditional kinetics among canonical quality-controlled transitions

Because inference on τ and t_onset_ was conditional on the prespecified physiological direction and fitting criteria, we next quantified entry into the kinetic analysis set. Retention under the prespecified fit-quality criterion (FitQC) was 87.7% for eye opening and 87.2% for eye closure in HC, compared with 58.7% for both conditions in MCI. Final canonical kinetic inclusion, meaning fulfillment of the complete conditional kinetic-analysis criteria, was 86.0% and 86.6% in HC versus 54.3% and 55.2% in MCI (Supplementary Table S1). Covariate-adjusted mixed-effects logistic models confirmed substantially lower odds of both FitQC and final canonical kinetic inclusion in MCI (all Holm-adjusted P<0.001; Supplementary Table S2 and Supplementary Fig. S1). A diagnostic breakdown of FitQC failure modes is shown in Supplementary Fig. S2. These findings define the conditional analysis set in which the kinetic estimates should be interpreted.

Among canonical quality-controlled transitions, no significant group difference in τ was detected during eye opening (MCI/HC ratio=1.056, 95% CI 0.880–1.268; Holm-adjusted P=1.000) or eye closure (ratio=0.951, 95% CI 0.805–1.123; Holm-adjusted P=1.000). Eye-opening t_onset_ was 0.107 s later in MCI (95% CI 0.033–0.182; Holm-adjusted P=0.020), whereas no significant eye-closure t_onset_ difference was detected (β=-0.024 s, 95% CI -0.159–0.110; Holm-adjusted P=1.000). Thus, within the retained canonical quality-controlled subset, the clearest detected between-group kinetic difference concerned transition onset (Table 2).

### Repetition-dependent alteration of transition onset

Given the preceding between-group differences, we next tested whether the transition measures showed group-dependent changes across the three repeated eye-state switches. No significant Group × Trial interaction was observed for LogEnergyRatio, transition success or the log-transformed time constant (LogTau) in either condition after Holm correction (Supplementary Table S3). In contrast, t_onset_ showed significant Group × Trial interactions for both eye opening (F(2,185.78)=8.138, P=0.00041; Holm-adjusted P=0.003) and eye closure (F(2,184.64)=8.222, P=0.00038; Holm-adjusted P=0.003) (Table 3 and Supplementary Table S3).

**Table 3.** Repetition-dependent changes in transition onset timing. Omnibus Group × Trial tests are shown together with model-based T3−T1 simple effects and participant-level T3−T1 sensitivity contrasts. Onset changes are in seconds relative to the refined ocular event; positive values indicate a later T3 onset. Paired kinetic observations were available for 49 HC/35 MCI participants during opening and 53 HC/35 MCI during closure. Kinetic comparisons are conditional on canonical quality-controlled transitions. Full omnibus models, continuous-Trial t_onset_ trends and ΔLogTau sensitivity results are reported in Supplementary Table S3.

| Condition | Group × Trial interaction | HC T3-T1 | MCI T3-T1 | T3-T1 sensitivity (MCI-HC) |
| --- | --- | --- | --- | --- |
| Open | F=8.138; P=0.00041; Holm P=0.003 | 0.239 s (0.136, 0.343); Holm P<0.001 | -0.076 s (-0.192, 0.039); Holm P=0.389 | -0.314 s (-0.463, -0.166); Holm P<0.001 |
| Close | F=8.222; P=0.00038; Holm P=0.003 | -0.409 s (-0.552, -0.267); Holm P<0.001 | 0.035 s (-0.128, 0.198); Holm P=1.000 | 0.337 s (0.140, 0.533); Holm P=0.001 |

The interaction reflected state-dependent temporal reorganization in HC that was not evident in MCI. By Trial 3, HC showed a 0.239-s delay relative to Trial 1 during eye opening (95% CI 0.136–0.343; Holm-adjusted P<0.001), whereas MCI showed no significant change (-0.076 s, 95% CI -0.192–0.039; Holm-adjusted P=0.389). During eye closure, HC showed a 0.409-s earlier onset at Trial 3 (95% CI -0.552 to - 0.267; Holm-adjusted P<0.001), whereas MCI again showed no significant change (0.035 s, 95% CI - 0.128–0.198; Holm-adjusted P=1.000) (Table 3).

Sensitivity analyses supported the same pattern. Treating Trial as a continuous variable yielded Group × Trial coefficients of -0.158 s/trial for eye opening (95% CI -0.235 to -0.081; Holm-adjusted P<0.001) and 0.222 s/trial for eye closure (95% CI 0.114–0.330; Holm-adjusted P<0.001) (Supplementary Table S3). In participant-level T3-T1 analyses, the adjusted MCI-minus-HC difference in the T3-T1 change in t_onset_ (Δt_onset_) was -0.314 s for eye opening (95% CI -0.463 to -0.166; Holm-adjusted P<0.001) and 0.337 s for eye closure (95% CI 0.140–0.533; Holm-adjusted P=0.001), whereas the T3-T1 change in LogTau (ΔLogTau) did not differ significantly between groups (Supplementary Table S3). The t_onset_ effects remained stable when the fitting threshold was varied across R^2^>0.30, 0.35 and 0.40 (Supplementary Table S4 and Supplementary Fig. S3).

### Associations with cognition in MCI

Within the MCI group, none of the six prespecified repetition-related EEG changes remained significantly associated with MoCA after Holm correction (Supplementary Table S5 and Supplementary Fig. S4). Close-eye ΔLogEnergyRatio showed a nominal Spearman correlation with MoCA (ρ=0.244, P=0.042; Holm-adjusted P=0.250), but the adjusted robust-regression estimate also did not reach significance (β=0.611, 95% CI -0.059–1.281; Holm-adjusted P=0.472). Open-eye ΔLogTau (ρ=-0.297, P=0.084) and Δt_onset_ (ρ=0.288, P=0.094) showed non-significant trends. Because paired T1/T3 kinetic measurements were available for only 35 MCI participants (Supplementary Tables S2 and S5), these analyses had a smaller effective sample size. Analyses using Δτ (τ at T3 minus τ at T1) instead of ΔLogTau did not materially change the conclusions.

## Discussion

This study demonstrates that repeated eye-state switching reveals two separable abnormalities in MCI: reduced expression of alpha-state transitions and disrupted repetition-dependent organization of transition onset. MCI participants retained the expected direction of alpha suppression after eye opening and alpha enhancement after eye closure, but both responses were smaller and transition success was lower. These findings extend prior primary studies showing abnormal resting EEG activity and impaired alpha reactivity in cognitive impairment [3,9,11] by emphasizing the dynamic range and reliability of state transitions rather than resting power or condition-averaged reactivity alone.

The bidirectional reduction in modulation is particularly informative. A weaker response in both directions is difficult to explain solely by a higher or lower baseline alpha level in one state. Instead, it is consistent with a narrower functional range over which posterior alpha activity can be reconfigured. This interpretation is compatible with experimental evidence that eye-state alpha and ongoing alpha fluctuations reflect changes in visual processing and internal-versus-external attentional engagement [6–8]. The markedly lower odds of canonical transitions in MCI further suggest that the abnormality concerns not only how strongly the system changes, but also whether the expected state change is reliably expressed. For this reason, the term “state-transition capacity” is used here as a descriptive construct integrating transition magnitude and transition probability, rather than implying a single underlying mechanism.

Among canonical quality-controlled transitions, no significant group difference in τ was detected in either eye-state condition, and the estimates were stable across the tested R^2^ thresholds. These results do not establish equivalence between groups; rather, the present data did not provide evidence for a marked shift in post-onset time scale within the retained kinetic subset. Because transition success and kinetic inclusion were substantially lower in MCI, inference on τ and t_onset_ is conditional on transitions that showed the expected physiological direction and met the fit-quality criteria.

The most distinctive finding was the repetition-dependent behavior of t_onset_. In HC, the timing relationship between alpha-state change and the observable eye movement was systematically reorganized across repetitions: t_onset_ became later relative to eye opening but earlier relative to eye closure. The opposite directions argue for state-specific temporal adaptation rather than a nonspecific practice effect. We interpret this pattern as flexible recalibration of the timing that links task instruction, state transition and overt eye behavior, a process that was markedly attenuated in MCI. This interpretation is consistent with literature linking alpha dynamics to shifts between internally and externally oriented processing [7,8] and with reports of altered temporal or network organization in MCI [12,16,18].

For eye opening, a plausible account is that alpha suppression is partly cue-driven rather than being triggered only by the physical act of opening the eyes. The verbal instruction can initiate attentional reorientation, task-set switching and motor preparation before measurable eyelid movement, processes that can modulate alpha activity [7,8,16]. With repeated instructions, HC participants may have initiated eye opening progressively sooner after the cue, while cue-related alpha suppression shifted less. If so, the ocular marker would move closer to the unrecorded instruction time and alpha suppression would appear progressively later relative to eye opening, even without a comparable delay relative to the cue. This provides a coherent behavioral-neural explanation for the HC repetition effect and suggests more flexible cue-brain-behavior coupling than in MCI. Because instruction times were not recorded, the present data cannot separate changes in neural latency from changes in behavioral response latency.

The eye-closure pattern points to a complementary form of adaptation. Closing the eyes withdraws visual input and requires re-establishment of an internally oriented, alpha-synchronized state; with repetition, the earlier t_onset_ in HC may therefore reflect more rapid re-engagement of the processes that restore posterior alpha activity [6–8]. The absence of a comparable shift in MCI is consistent with reduced flexibility in reorganizing brain state across repeated transitions. Importantly, the opposite direction from eye opening suggests that repetition did not simply speed all motor responses, but altered the temporal coupling between neural state change and behavior in a state-dependent manner. The present design cannot identify the specific neural pathway responsible, and future studies with time-stamped auditory cues and independent eyelid tracking can test whether cue-to-alpha and cue-to-movement latencies change separately across repetitions.

The lack of robust associations with MoCA may indicate that these transition measures capture a physiological dimension of dynamic state regulation that is not well represented by a brief global cognitive score [25]. Transition timing may be more closely related to specific processes such as attentional switching, executive control, processing speed or task preparation [16,18]. The smaller effective sample for paired kinetic changes (35 MCI participants) also limited sensitivity; domain-specific cognitive measures and larger longitudinal cohorts may therefore be more informative for testing clinical relevance.

Several methodological features strengthen the study. Frontal ocular artifacts were used to refine conventional manual EEG markers, allowing the classic eye-opening/eye-closure response to be analyzed as a time-resolved process. Transition magnitude, canonical-transition probability, t_onset_ and τ were treated as distinct dimensions, avoiding over-reliance on a single summary measure. Repeated trials were modeled with participant-specific random effects [26], and the principal t_onset_ finding was supported by categorical-Trial, linear-trend, T3-T1 and R^2^-threshold sensitivity analyses.

The study also has limitations. It was cross-sectional and single-center, so the observed EEG phenotype cannot be assumed to predict future decline or conversion to dementia. The 19-channel clinical EEG montage has limited spatial resolution, and the 200-Hz source sampling constrains event-time precision to 5 ms despite the finer interpolation grid used for numerical candidate localization. Eye-state timing was estimated from Fp1/Fp2 artifacts rather than an independent electrooculogram, and the exact timing of spoken instructions was not recorded; auditory processing, cognitive preparation, eyelid movement and visual input therefore cannot be separated. ICA rejection, event confirmation and part of the occipital-channel quality assessment involved manual review, which may limit reproducibility. The pre-fit direction screen permuted individual time samples between the pre- and post-event windows and therefore did not preserve within-window temporal autocorrelation; it was used as an operational screening step before fitting rather than as the primary group-level inferential test. The mono-exponential model provides a parsimonious description of the dominant transition but cannot capture all multiphasic or non-exponential dynamics. Finally, the lower kinetic-analysis inclusion rate in MCI means that τ and t_onset_ describe a conditional subset of canonical quality-controlled transitions. Independent cohorts and longitudinal follow-up will be needed to establish robustness and clinical significance.

In conclusion, MCI was associated with weaker and less reliable bidirectional alpha-state transitions during repeated eye opening and eye closure. Among canonical quality-controlled transitions, no significant group difference in τ was detected, whereas transition-onset timing showed a more prominent group-dependent pattern. Healthy controls exhibited state-dependent temporal reorganization of t_onset_ across repeated trials, while MCI participants did not show a comparable change. Repeated eye-state EEG may therefore provide a simple physiological paradigm for probing dynamic brain-state regulation in MCI, with kinetic inferences restricted to successfully expressed and quality-controlled canonical transitions.

## Methods

### Study design and participants

This cross-sectional EEG study was conducted in the Department of Neurology at the Affiliated Hospital of Jiangsu University. The protocol was approved by the institutional ethics committee (KY2024K1102), and written informed consent was obtained from all participants or their legal representatives. All procedures were performed in accordance with the Declaration of Helsinki and relevant institutional guidelines and regulations. Participants were recruited from neurology and cognitive clinics and from health-examination populations. HC participants had no known neurodegenerative disease, cerebrovascular disease, epilepsy or other neurological disorder expected to substantially affect cognition or EEG and were able to understand and complete the eye-state instructions. MCI was diagnosed by neurologists from clinical history, subjective cognitive concerns, objective cognitive assessment and preserved everyday function, following established Petersen criteria [1,2]. MoCA supported cognitive assessment, with one point added for participants with ≤12 years of education [25]. Clinical variables included age, sex, education, MoCA, MMSE and CDR; CDR was used for clinical characterization rather than as a continuous model covariate.

### EEG acquisition and eye-state paradigm

EEG was recorded with a Neurofax EEG-1200C system (Nihon Kohden) from 19 scalp electrodes positioned according to the international 10–20 system [28], with A1/A2 earlobe references and electrode impedances <5 kΩ. The EDF data used for offline analysis had a sampling frequency of 200 Hz, corresponding to a 5-ms interval between original samples. Participants first rested comfortably with their eyes closed for at least 20 s and then completed three consecutive eye-state cycles following clear verbal instructions. Each cycle comprised approximately 10 s of eye opening followed by approximately 10 s of eye closure; after the third opening, participants closed their eyes again and remained in that state for at least 20 s. Manual event markers were inserted when the examiner observed the eye movement, but these markers were not treated as the final event times because of possible operator-to-eyelid timing offsets (Fig. 1a).

### EEG preprocessing and event localization

Offline analyses were performed in MATLAB R2021a (MathWorks, Natick, MA, USA) using custom code. Lateral scalp channels were referenced to the corresponding earlobe electrode, whereas midline channels followed the prespecified average-reference convention. Signals were processed with zero-phase forward-and-reverse Butterworth filtering (0.5–70 Hz) and a 49–51 Hz notch filter to reduce 50-Hz line noise. The analyzed continuous segment included baseline before the first target event and recovery after the final event, and filtering edge transients were excluded. After event timing had been established, extended Infomax independent component analysis was applied [29]. Components dominated by ocular, blink or obvious muscle activity were removed after visual inspection, and the cleaned EEG was reconstructed from the remaining components. To refine eye-state timing, Fp1 and Fp2 were averaged as an EOG-like signal and smoothed. For numerical candidate localization only, this signal was cubic-spline interpolated from 200 Hz to 20 kHz. First- and second-derivative features within approximately ±2 s of each manual marker generated candidate eye-opening and eye-closure events. Candidates were then manually confirmed from the frontal ocular-proxy waveform and its derivative relative to the original marker (Fig. 1b); occipital EEG was not used to determine the event time. The interpolation provided a smooth numerical grid but did not increase the physiological temporal resolution beyond the 200-Hz source signal. The refined ocular event was used as t=0 for subsequent event-locked analyses.

### Alpha-energy extraction and transition measures

The primary analysis used occipital O1/O2 activity in the 8–12 Hz alpha band. For each refined event, the program initially requested up to approximately 4 s of data before and after the event and adaptively truncated the window when necessary to avoid overlap with adjacent eye-state events. Alpha-band signals were Hilbert transformed [30], and instantaneous energy was defined as the squared analytic amplitude. O1 and O2 were reviewed at the trial level; channels with obvious artifact contamination were excluded, and the remaining usable occipital energy traces were averaged to obtain a representative alpha-energy trajectory for that trial.

Before kinetic fitting, pre- and post-event 4-s alpha-energy windows were compared using a 1,000-iteration two-sided sample-label permutation procedure. Within each trial, individual time samples from the two windows were pooled and randomly reassigned to pseudo-pre and pseudo-post groups of the original sizes; the difference in medians was recomputed for each permutation. The observed median difference was also standardized by a pooled median-absolute-deviation-derived scale. A trial was classified as increasing (k_dir_=+1) when P<0.05 and the robust effect size was >0.5, decreasing (k_dir_=-1) when P<0.05 and the effect size was <-0.5, and otherwise as showing no clear change; trials in the last category were not kinetically fitted. Transition magnitude was summarized by EnergyRatio, the ratio of mean post-event to mean pre-event alpha energy, with MATLAB’s positive machine-epsilon constant (ε) added to the denominator to prevent division by zero. LogEnergyRatio was the natural logarithm of EnergyRatio. LogEnergyRatio=0 corresponds to no net energy change, with negative values indicating lower post-event energy and positive values higher post-event energy. TransitionSuccess was the binary indicator of the expected physiological direction, defined as EnergyRatio<1 for eye opening and EnergyRatio>1 for eye closure.

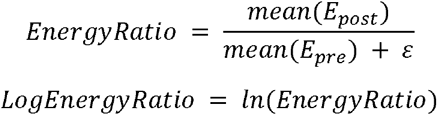

### Kinetic modeling and quality control

Trials with a clear energy change were fitted by nonlinear least squares using the mono-exponential model shown below. The directional coefficient k_dir_ was fixed by the preceding classification (+1 for increasing and -1 for decreasing), while A□ and τ were constrained to positive values. Here, C is baseline energy, A□ is transition amplitude, H is the Heaviside step function, t_onset_ is the fitted transition onset, and τ>0 is the fitted post-onset time constant. FitQC required R^2^>0.35, τ>0.03 s, and finite τ and t_onset_ values. Conditional kinetic analyses additionally required the expected physiological direction: eye opening had to satisfy EnergyRatio<1 and k_dir_=-1, whereas eye closure had to satisfy EnergyRatio>1 and k_dir_=+1. These trials are referred to as canonical quality-controlled transitions. Because τ was right-skewed, analyses used LogTau=ln(τ); exp(β) therefore represents the MCI/HC ratio on the original τ scale. Repetition-related change was defined as ΔLogTau=LogTau_T3_−LogTau_T1_. The untransformed sensitivity measure was defined as Δτ=τ_T3_−τ_T1_. R^2^ thresholds of >0.30 and >0.40 were also examined to test threshold sensitivity (Supplementary Table S4 and Supplementary Fig. S3).

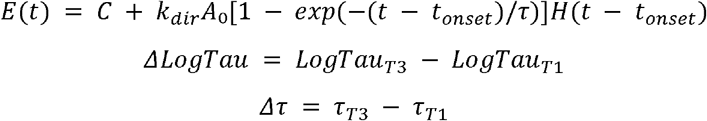

### Data quality and analysis hierarchy

TransitionSuccess and FitQC were treated as conceptually distinct. TransitionSuccess describes whether alpha energy changed in the expected physiological direction, whereas FitQC describes whether the fitted kinetic parameters met the prespecified technical criteria. LogEnergyRatio and TransitionSuccess formed the first-stage transition-capacity analyses and used all available Trial 1–3 transitions; τ and t_onset_ formed conditional second-stage analyses restricted to canonical quality-controlled transitions. Because of occasional acquisition or marker errors, seven participants contributed two recorded cycles and one participant contributed one cycle, while two participants had an additional fourth cycle. Primary analyses retained all available Trials 1–3 and excluded Trial 4; participants were not excluded solely because the recorded cycle count differed from three. Analyses of T3−T1 change required the relevant observations at both trials. Supplementary mixed-effects logistic models quantified group differences in FitQC and final canonical kinetic inclusion (Supplementary Table S2 and Supplementary Fig. S1), and R^2^-threshold sensitivity analyses repeated the conditional kinetic and Group × Trial models at R^2^>0.30, >0.35 and >0.40 (Supplementary Table S4 and Supplementary Fig. S3).

### Statistical analysis

All statistical analyses were performed in MATLAB R2021a and were two-sided, with P<0.05 considered significant. Continuous demographic variables were compared with Mann-Whitney U tests and sex with Fisher’s exact test. For paradigm validation, LogEnergyRatio was first averaged across the three trials within each participant; eye-opening and eye-closure participant means were then tested against zero using Wilcoxon signed-rank tests, and the proportion of participants showing the expected direction was reported. Eye opening and eye closure were modeled separately for trial-level analyses. LogEnergyRatio, LogTau and t_onset_ were analyzed with linear mixed-effects models, and TransitionSuccess with a binomial generalized linear mixed-effects model. For primary between-group models, Trial was entered as a linear covariate coded 0, 1 and 2; models also included group, age, sex and education as fixed effects and participant as a random intercept, with HC as the reference group [26]. For the primary between-group comparisons, Holm adjustment was applied separately within three prespecified families: LogEnergyRatio across eye opening and eye closure; TransitionSuccess across eye opening and eye closure; and conditional kinetic outcomes comprising LogTau and t_onset_ in each eye-state condition. For repetition analyses, Trial was treated primarily as a categorical variable with T1 as the reference, and the T2 and T3 Group × Trial coefficients were tested jointly. For omnibus F statistics, the two values in parentheses denote the numerator and denominator degrees of freedom, respectively. The omnibus Group × Trial tests across the four outcomes and two eye-state conditions were Holm-adjusted as a separate family [31]. Significant t_onset_ interactions were followed by within-group T2-T1 and T3-T1 contrasts with within-condition Holm adjustment. Sensitivity analyses treated Trial as a continuous variable and compared participant-level T3-T1 changes using robust regression adjusted for age, sex and education (Supplementary Table S3). Within MCI, repetition-related EEG changes were related to MoCA using Spearman correlations and adjusted robust regression; the six EEG measures were Holm-corrected as a separate family, and Δτ was evaluated as an untransformed sensitivity analysis (Supplementary Table S5).

### Use of generative artificial intelligence (AI)

Generative artificial intelligence was used to assist with English-language rewriting and structural editing of the manuscript. It was not used to generate or alter study data or to perform the reported statistical analyses. All AI-assisted text was reviewed and revised by the authors, who remain responsible for the accuracy, interpretation and integrity of the manuscript.

## Data Availability

De-identified data supporting the findings of this study are available from the corresponding author upon reasonable request, subject to applicable institutional and ethical requirements for protection of participant confidentiality. Requests should be directed to Ming Yu.

## Acknowledgements

The authors thank all participants for their participation in this study.

## Author contributions

Xuan Wang: Conceptualization, Data curation, Investigation, Writing – original draft. Ying Zhu: Project administration, Supervision, Writing – review & editing. Jian Zhang: Data curation, Investigation. Zheng Zheng: Data curation, Investigation. Ming Yu: Conceptualization, Methodology, Project administration, Supervision, Writing – review & editing. All authors reviewed and approved the final manuscript.

## Code availability

The custom MATLAB code used for EEG preprocessing, eye-state event localization, kinetic fitting and statistical analyses is available from the corresponding author, Ming Yu, upon reasonable request.

## References

1. Petersen, R. C. Mild cognitive impairment. N. Engl. J. Med. 364, 2227–2234 (2011).

2. Albert, M. S. et al. The diagnosis of mild cognitive impairment due to Alzheimer’s disease: recommendations from the National Institute on Aging-Alzheimer’s Association workgroups on diagnostic guidelines for Alzheimer’s disease. Alzheimers Dement. 7, 270–279 (2011).

3. Jiang, Y. et al. Abnormal resting-state EEG neural oscillations and functional connectivity in mild cognitive impairment. Front. Aging Neurosci. 17, 1640966 (2025).

4. Kim, S.-K. et al. Electroencephalography-based classification of Alzheimer’s disease spectrum during computer-based cognitive testing. Sci. Rep. 14, 5252 (2024).

5. Jiang, Y. et al. Optimizing electrode configurations for EEG mild cognitive impairment detection. Sci. Rep. 15, 578 (2025).

6. Hohaia, W., Saurels, B. W., Johnston, A., Yarrow, K. & Arnold, D. H. Occipital alpha-band brain waves when the eyes are closed are shaped by ongoing visual processes. Sci. Rep. 12, 1194 (2022).

7. Zhang, Y. et al. Shared oscillatory mechanisms of alpha-band activity in prefrontal regions in eyes open and closed state using a portable EEG acquisition device. Sci. Rep. 14, 26719 (2024).

8. Kasten, F. H., Busson, Q. & Zoefel, B. Opposing neural processing modes alternate rhythmically during sustained auditory attention. Commun. Biol. 7, 1125 (2024).

9. Babiloni, C. et al. Reactivity of cortical alpha rhythms to eye opening in mild cognitive impairment and Alzheimer’s disease: an EEG study. J. Alzheimers Dis. 22, 1047–1064 (2010).

10. Chae, S. et al. Decreased alpha reactivity from eyes-closed to eyes-open in non-demented older adults with Alzheimer’s disease: a combined EEG and [18F]florbetaben PET study. J. Alzheimers Dis. 77, 1681–1692 (2020).

11. Babiloni, C. et al. Resting state electroencephalographic alpha rhythms are sensitive to Alzheimer’s disease mild cognitive impairment progression at a 6-month follow-up. Neurobiol. Aging 137, 19–37 (2024).

12. Wu, X. et al. Unveiling neural activity changes in mild cognitive impairment using microstate analysis and machine learning. J. Alzheimers Dis. 103, 735–748 (2025).

13. Kim, J.-G. et al. Differentiating amnestic from non-amnestic mild cognitive impairment subtypes using graph theoretical measures of electroencephalography. Sci. Rep. 12, 6219 (2022).

14. Carpi, M. et al. Periodic and aperiodic electroencephalographic rhythms during vigilance transitions in Alzheimer’s disease mild cognitive impairment. Ann. N. Y. Acad. Sci. 1555, e70194 (2026).

15. Salamone, E. M. et al. Abnormal electroencephalographic rhythms from quiet wakefulness to light sleep in Alzheimer’s disease patients with mild cognitive impairment. Clin. Neurophysiol. 171, 164–181 (2025).

16. Fodor, Z. et al. EEG alpha and beta band functional connectivity and network structure mark hub overload in mild cognitive impairment during memory maintenance. Front. Aging Neurosci. 13, 680200 (2021).

17. Jiang, Y., Zhang, X., Guo, Z. & Jiang, N. Altered EEG theta and alpha band functional connectivity in mild cognitive impairment during working memory coding. IEEE Trans. Neural Syst. Rehabil. Eng. 32, 2845–2853 (2024).

18. Wang, L. et al. Cerebral dominance representation of directed connectivity within and between left-right hemispheres and frontal-posterior lobes in mild cognitive impairment. Cereb. Cortex 33, 11279–11286 (2023).

19. Hamilton, C. A. et al. Slowing on quantitative EEG is associated with transition to dementia in mild cognitive impairment. Int. Psychogeriatr. 33, 1321–1325 (2021).

20. Chu, K.-T. et al. A holo-spectral EEG analysis provides an early detection of cognitive decline and predicts the progression to Alzheimer’s disease. Front. Aging Neurosci. 15, 1195424 (2023).

21. Hasoon, J. et al. EEG functional connectivity differences predict future conversion to dementia in mild cognitive impairment with Lewy body or Alzheimer disease. Int. J. Geriatr. Psychiatry 39, e6138 (2024).

22. Ge, Y. et al. An EEG-based framework for automated discrimination of conversion to Alzheimer’s disease in patients with amnestic mild cognitive impairment: an 18-month longitudinal study. Front. Aging Neurosci. 16, 1470836 (2025).

23. Aoki, Y. et al. EEG resting-state networks in Alzheimer’s disease associated with clinical symptoms. Sci. Rep. 13, 3964 (2023).

24. Aljalal, M., Aldosari, S. A., Molinas, M. & Alturki, F. A. Selecting EEG channels and features using multi-objective optimization for accurate MCI detection: validation using leave-one-subject-out strategy. Sci. Rep. 14, 12483 (2024).

25. Nasreddine, Z. S. et al. The Montreal Cognitive Assessment, MoCA: a brief screening tool for mild cognitive impairment. J. Am. Geriatr. Soc. 53, 695–699 (2005).

26. Laird, N. M. & Ware, J. H. Random-effects models for longitudinal data. Biometrics 38, 963–974 (1982).

27. Peltola, M. E. et al. Routine and sleep EEG: minimum recording standards of the International Federation of Clinical Neurophysiology and the International League Against Epilepsy. Epilepsia 64, 602–618 (2023).

28. Jasper, H. H. The ten-twenty electrode system of the International Federation. Electroencephalogr. Clin. Neurophysiol. 10, 371–375 (1958).

29. Lee, T. W., Girolami, M. & Sejnowski, T. J. Independent component analysis using an extended infomax algorithm for mixed subgaussian and supergaussian sources. Neural Comput. 11, 417–441 (1999).

30. Le Van Quyen, M. et al. Comparison of Hilbert transform and wavelet methods for the analysis of neuronal synchrony. J. Neurosci. Methods 111, 83–98 (2001).

31. Holm, S. A simple sequentially rejective multiple test procedure. Scand. J. Stat. 6, 65–70 (1979).

