## Supplementary Information for "Attenuated alpha state transitions and disrupted repetition-dependent temporal adaptation in mild cognitive impairment"

Supplementary Results, Tables and Figure Legends

**Supplementary Results**

### Supplementary Result 1. Paradigm validation and data-retention overview

Across all participants, mean LogEnergyRatio was negative after eye opening (-1.202 ± 0.964) and positive after eye closure (1.313 ± 1.065), with 90.5% and 92.7% of participants showing the expected direction, respectively (both P < 0.001). At the trial level, 344/402 (85.6%) eye-opening transitions and 352/402 (87.6%) eye-closure transitions showed transition success. Group-specific transition success, FitQC retention and canonical kinetic inclusion are summarized in Supplementary Table S1.

**Supplementary Table S1 | Paradigm validation and data-retention summary.**

**A. Subject-level paradigm validation**

| **Population** | **Condition** | **N** | **Mean ± SD LogER** | **Median [IQR] LogER** | **Expected direction (%)** | **P versus 0** |
| --- | --- | --- | --- | --- | --- | --- |
| All | Open | 137 | -1.202 ± 0.964 | -1.150 [-1.821, -0.410] | 90.5 | <0.001 |
| All | Close | 137 | 1.313 ± 1.065 | 1.078 [0.529, 2.084] | 92.7 | <0.001 |
| HC | Open | 60 | -1.647 ± 0.718 | -1.641 [-2.079, -1.156] | 100.0 | <0.001 |
| HC | Close | 60 | 1.833 ± 0.978 | 1.733 [1.141, 2.489] | 98.3 | <0.001 |
| MCI | Open | 77 | -0.855 ± 0.991 | -0.601 [-1.401, -0.100] | 83.1 | <0.001 |
| MCI | Close | 77 | 0.908 ± 0.954 | 0.727 [0.280, 1.178] | 88.3 | <0.001 |

LogER, LogEnergyRatio; IQR, interquartile range; HC, healthy controls; MCI, mild cognitive impairment.

**B. Trial-level transition and quality-control summary**

| **Group** | **Condition** | **Trial rows** | **Transition success n (%)** | **FitQC n (%)** | **Canonical kinetic n (%)** |
| --- | --- | --- | --- | --- | --- |
| All | Open | 402 | 344 (85.6) | 288 (71.6) | 275 (68.4) |
| All | Close | 402 | 352 (87.6) | 287 (71.4) | 278 (69.2) |
| HC | Open | 179 | 170 (95.0) | 157 (87.7) | 154 (86.0) |
| MCI | Open | 223 | 174 (78.0) | 131 (58.7) | 121 (54.3) |
| HC | Close | 179 | 170 (95.0) | 156 (87.2) | 155 (86.6) |
| MCI | Close | 223 | 182 (81.6) | 131 (58.7) | 123 (55.2) |

Transition success denotes the expected direction of alpha-energy change. Canonical kinetic inclusion required TransitionSuccess, FitQC and the expected model direction.

**C. Trial-level descriptive EnergyRatio medians**

| **Group** | **Condition** | **Trial rows** | **Median EnergyRatio** |
| --- | --- | --- | --- |
| HC | Open | 179 | 0.178 |
| MCI | Open | 223 | 0.536 |
| HC | Close | 179 | 5.88 |
| MCI | Close | 223 | 1.91 |

EnergyRatio medians are descriptive summaries across the available trial-level transitions in each group and condition.

### Supplementary Result 2. Kinetic fit quality and inclusion in conditional kinetic analyses

Under the primary fit-quality criterion of R² > 0.35, kinetic-fit retention differed substantially between HC and MCI. During eye opening, 87.7% of HC trials met FitQC compared with 58.7% of MCI trials; during eye closure, the corresponding proportions were 87.2% and 58.7%.

In mixed-effects logistic models adjusted for age, sex, years of education and Trial, with participant included as a random intercept, MCI was associated with markedly lower odds of meeting FitQC. The adjusted MCI-versus-HC odds ratio was 0.174 (95% CI 0.085–0.356; Holm-adjusted P < 0.001) during eye opening and 0.187 (95% CI 0.094–0.372; Holm-adjusted P < 0.001) during eye closure.

We next examined the final set of trials eligible for canonical conditional-kinetic analyses, defined as trials with the expected direction of alpha-power change, acceptable fit quality and the expected model direction. Canonical kinetic inclusion was achieved in 86.0% and 86.6% of HC trials during eye opening and eye closure, respectively, compared with 54.3% and 55.2% of MCI trials. After covariate adjustment, the odds ratio for canonical kinetic inclusion in MCI versus HC was 0.164 during both eye opening (95% CI 0.081–0.329; Holm-adjusted P < 0.001) and eye closure (95% CI 0.081–0.331; Holm-adjusted P < 0.001).

At the participant level, all 60 HC participants had at least one canonical kinetic trial during eye opening and 58/60 (96.7%) did so during eye closure. The corresponding proportions in MCI were 55/77 (71.4%) and 58/77 (75.3%). Availability was further reduced for paired T1 and T3 change analyses: valid T1+T3 measurements were available for 49/60 (81.7%) HC and 35/77 (45.5%) MCI participants during eye opening, and for 53/60 (88.3%) HC and 35/77 (45.5%) MCI participants during eye closure. Accordingly, analyses of ΔLogTau and Δt_onset_ are conditional on participants who produced canonical, quality-controlled transitions at the required trials.

**Supplementary Table S2 | FitQC and probability of inclusion in conditional kinetic analyses.**

**A. Adjusted mixed-effects logistic models**

| **Condition** | **Outcome** | **OR, MCI vs HC** | **95% CI** | **Holm P** |
| --- | --- | --- | --- | --- |
| Open | FitQC | 0.174 | 0.085, 0.356 | <0.001 |
| Close | FitQC | 0.187 | 0.094, 0.372 | <0.001 |
| Open | Canonical kinetic inclusion | 0.164 | 0.081, 0.329 | <0.001 |
| Close | Canonical kinetic inclusion | 0.164 | 0.081, 0.331 | <0.001 |

Models were adjusted for age, sex, years of education and Trial, with a participant-specific random intercept. OR, odds ratio; CI, confidence interval.

**B. Participant-level availability**

| **Condition** | **Group** | **Total N** | **≥1 canonical fit n (%)** | **Valid T1+T3 pair n (%)** |
| --- | --- | --- | --- | --- |
| Open | HC | 60 | 60 (100.0) | 49 (81.7) |
| Open | MCI | 77 | 55 (71.4) | 35 (45.5) |
| Close | HC | 60 | 58 (96.7) | 53 (88.3) |
| Close | MCI | 77 | 58 (75.3) | 35 (45.5) |

A valid T1+T3 pair was required for participant-level change analyses. HC, healthy controls; MCI, mild cognitive impairment.

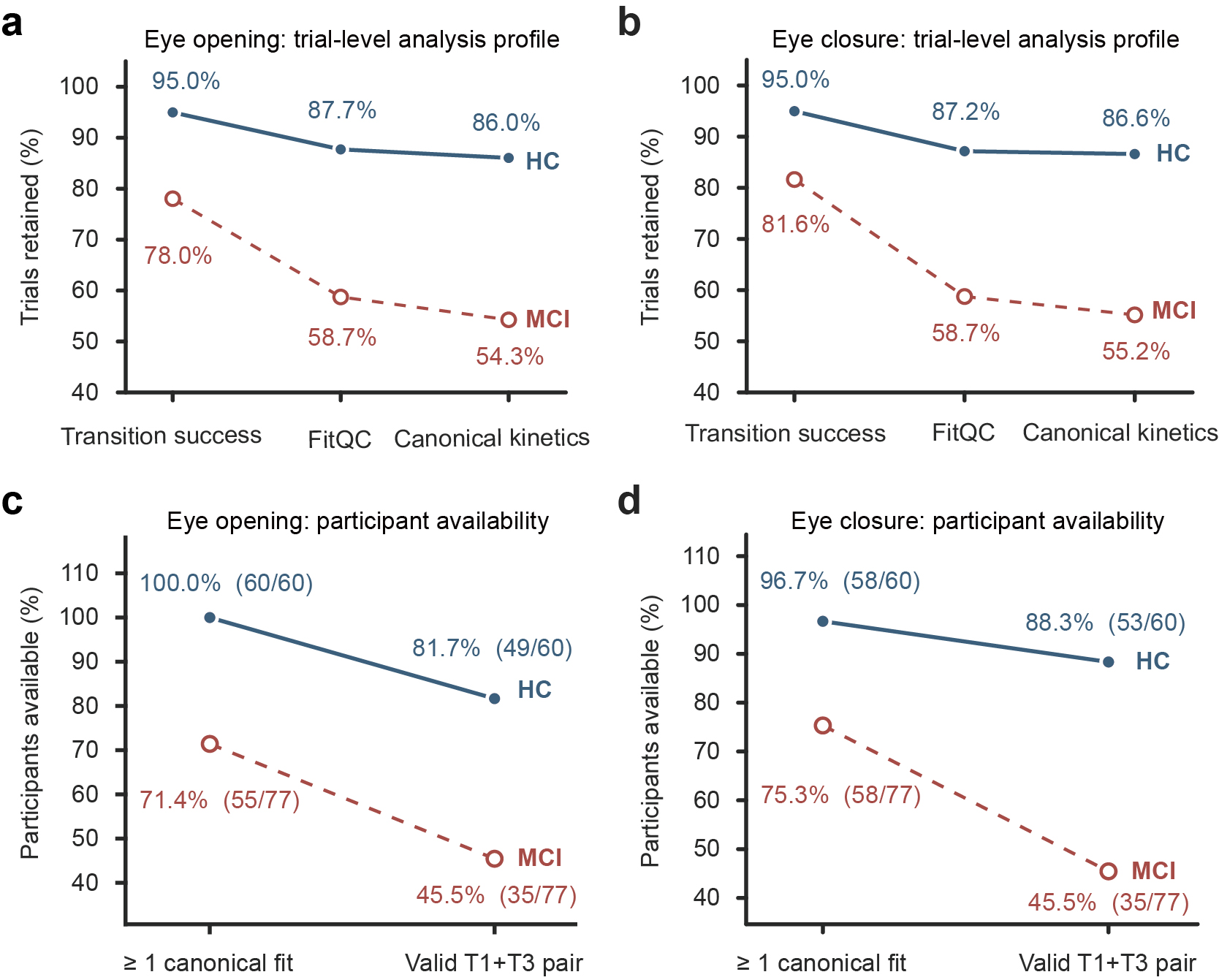

**Supplementary Figure S1 | Trial-level retention and participant availability for conditional kinetic analyses.**

**a,b**, Trial-level percentages meeting TransitionSuccess, FitQC and final canonical kinetic inclusion for eye opening (a) and closure (b). Denominators are 179 HC and 223 MCI trial rows per condition. **c,d**, Participant availability for at least one canonical fit and for a valid T1+T3 pair during opening (c) and closure (d); group denominators are 60 HC and 77 MCI. Values show percentages and n/N where applicable. HC, healthy controls; MCI, mild cognitive impairment.

#### Diagnostic profile of kinetic-fit failures

During eye opening, 22/179 (12.3%) HC trials and 92/223 (41.3%) MCI trials failed to meet the prespecified FitQC criterion. During eye closure, FitQC failure occurred in 23/179 (12.8%) HC trials and 92/223 (41.3%) MCI trials.

The failure components must be distinguished from one another (Supplementary Fig. S2). In eye opening, technical invalidity affected 12.3% of HC and 38.1% of MCI trial rows; among technically valid fits, the additional R² ≤ 0.35 category accounted for 0.0% and 3.1% of all HC and MCI trial rows, respectively. During eye closure, the corresponding percentages were 11.7% and 37.7% for technical invalidity and 1.1% and 3.6% for R² ≤ 0.35. No technically valid fit was classified as failing the τ ≤ 0.03 s criterion in the plotted diagnostic summary. Overall FitQC failure remained 12.3% and 41.3% in eye opening and 12.8% and 41.3% in eye closure (HC and MCI, respectively). These percentages use all trial rows as denominators and should not be summed across categories; in particular, an invalid fit must not automatically be labeled an R²-threshold failure.

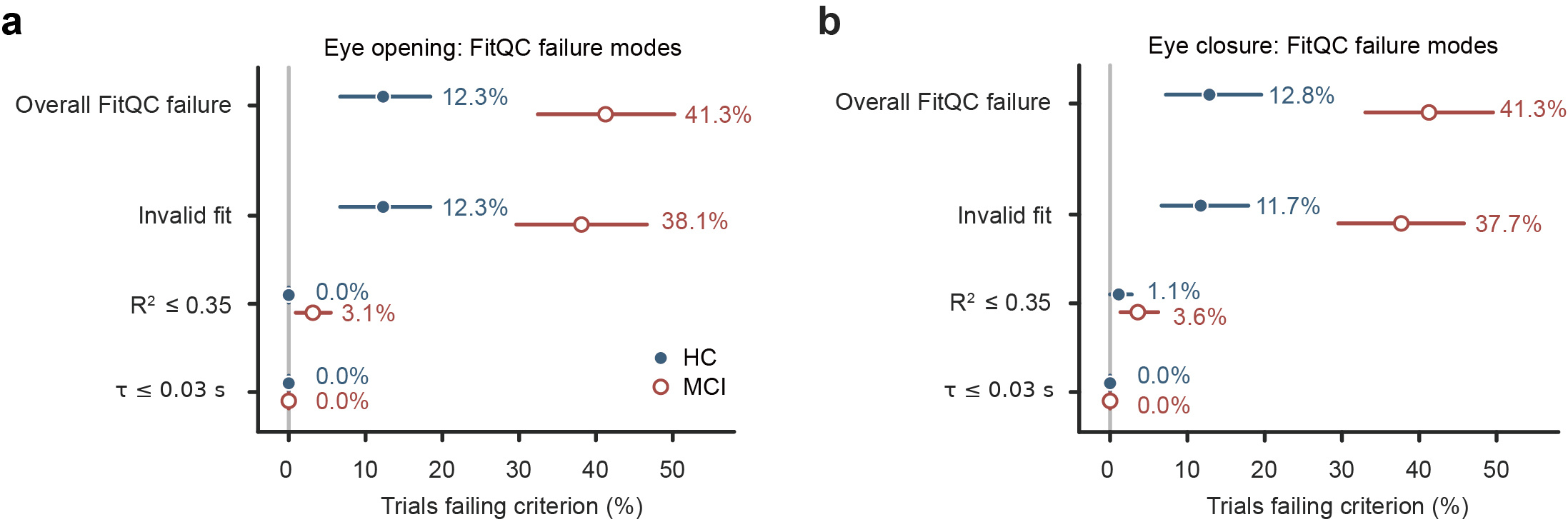

**Supplementary Figure S2 | Trial-level FitQC failure modes during eye-opening and eye-closure transitions.**

**a,b**, Components of FitQC failure for eye opening (a) and closure (b). Points show observed percentages and horizontal bars participant-cluster bootstrap 95% confidence intervals (5,000 resamples); denominators are 179 HC and 223 MCI trial rows per condition. Invalid fit denotes the absence of a technically valid kinetic estimate; the R²≤0.35 and τ≤0.03 s categories are evaluated among technically valid fits. HC, healthy controls; MCI, mild cognitive impairment.

### Supplementary Result 3. Repetition-effect models and sensitivity analyses

The primary categorical-Trial models are summarized in Supplementary Table S3. To test the average linear repetition trend, Trial was additionally coded as a continuous variable (0, 1 and 2).

In contrast, t_onset_ showed significant group-dependent linear trends in opposite directions across the two conditions. The Group × Trial interaction coefficient was -0.158 s/trial during eye opening (95% CI -0.235 to -0.081; Holm-adjusted P < 0.001) and 0.222 s/trial during eye closure (95% CI 0.114–0.330; Holm-adjusted P < 0.001).

A participant-level T3−T1 sensitivity analysis restricted to participants with valid kinetic data at both trials provided convergent evidence. No significant between-group difference was observed for ΔLogTau during either eye opening or eye closure, whereas Δt_onset_ differed significantly between groups in both conditions, consistent with the primary categorical-Trial analysis.

**Supplementary Table S3 | Group × Trial models and T3−T1 sensitivity analyses.**

**A. Omnibus Group × Trial tests in the primary models**

| **Condition** | **Outcome** | **F** | **P omnibus** | **Holm P** |
| --- | --- | --- | --- | --- |
| Open | LogEnergyRatio | 0.645 | 0.525 | 1.000 |
| Close | LogEnergyRatio | 0.673 | 0.511 | 1.000 |
| Open | TransitionSuccess | 0.012 | 0.988 | 1.000 |
| Close | TransitionSuccess | 1.110 | 0.331 | 1.000 |
| Open | LogTau | 0.292 | 0.747 | 1.000 |
| Close | LogTau | 0.160 | 0.853 | 1.000 |
| Open | t_onset_ | 8.138 | <0.001 | 0.003 |
| Close | t_onset_ | 8.222 | <0.001 | 0.003 |

Holm correction was applied across the prespecified omnibus Group × Trial tests.

**B. T3−T1 sensitivity analysis**

| **Outcome** | **Condition** | **N, HC / MCI** | **Median HC** | **Median MCI** | **Adjusted β** | **95% CI** | **Holm P** |
| --- | --- | --- | --- | --- | --- | --- | --- |
| ΔLogTau | Open | 49 / 35 | 0.079 | -0.137 | -0.203 | -0.671, 0.266 | 0.798 |
| ΔLogTau | Close | 53 / 35 | 0.046 | -0.086 | -0.107 | -0.414, 0.199 | 0.798 |
| Δt_onset_, s | Open | 49 / 35 | 0.232 | -0.066 | -0.314 | -0.463, -0.166 | <0.001 |
| Δt_onset_, s | Close | 53 / 35 | -0.238 | -0.022 | 0.337 | 0.140, 0.533 | 0.001 |

Changes were defined as T3−T1. HC, healthy controls; MCI, mild cognitive impairment; CI, confidence interval.

**C. Continuous-Trial sensitivity analysis for t_onset_**

| **Condition** | **Linear Group × Trial β, s/trial** | **95% CI** | **Holm P** |
| --- | --- | --- | --- |
| Open | -0.158 | -0.235, -0.081 | <0.001 |
| Close | 0.222 | 0.114, 0.330 | <0.001 |

Trial was coded 0, 1 and 2. β denotes the MCI-versus-HC difference in the linear repetition trend for t_onset_.

### Supplementary Result 4. Sensitivity of kinetic findings to the curve-fitting quality threshold

To determine whether the kinetic findings depended on the prespecified curve-fitting quality threshold, we repeated the conditional kinetic analyses and Group × Trial analyses after redefining FitQC using R² thresholds of 0.30 and 0.40, in addition to the primary threshold of R² > 0.35. All definitions continued to require τ > 0.03 s and technically valid kinetic estimates.

Changing the R² threshold produced only minor differences in data retention. During eye opening, the FitQC retention rate in HC was 87.7% at R² thresholds of 0.30, 0.35 and 0.40; the corresponding rates in MCI were 59.2%, 58.7% and 58.7%. During eye closure, the HC retention rate was approximately 87.2% at all three thresholds, whereas the MCI rates were 59.6%, 58.7% and 58.3%, respectively.

The principal kinetic results were stable across thresholds. During eye opening, the between-group difference in LogTau remained non-significant at R² thresholds of 0.30, 0.35 and 0.40. By contrast, the adjusted MCI-versus-HC effect on t_onset_ remained directionally consistent and statistically significant, with β values of 0.112, 0.107 and 0.107 s (P = 0.0033, 0.0051 and 0.0051, respectively). The between-group LogTau difference during eye closure was likewise non-significant across all thresholds.

The Group × Trial findings were similarly robust. For t_onset_, the omnibus interaction P values at R² thresholds of 0.30, 0.35 and 0.40 were 0.00027, 0.00041 and 0.00041 during eye opening, and 0.00020, 0.00038 and 0.00028 during eye closure. In contrast, no significant Group × Trial interaction for LogTau was observed at any threshold in either condition.

The continuous-trial models yielded the same pattern. The linear Group × Trial coefficient for t_onset_ was -0.162, -0.158 and -0.158 s/trial during eye opening and 0.233, 0.222 and 0.228 s/trial during eye closure at R² thresholds of 0.30, 0.35 and 0.40, respectively; all corresponding interaction tests had P < 0.001. Linear Group × Trial interactions for LogTau were non-significant at all thresholds. These findings indicate that the abnormal repetition-dependent pattern of t_onset_ was not driven by the specific choice of R² = 0.35, while the null findings for τ were also stable across the tested thresholds.

**Supplementary Table S4 | Sensitivity of data retention and kinetic results to the R² threshold.**

**A. FitQC retention by R² threshold**

| **R² threshold** | **Condition** | **HC, n (%)** | **MCI, n (%)** |
| --- | --- | --- | --- |
| 0.30 | Open | 157 (87.7) | 132 (59.2) |
| 0.30 | Close | 156 (87.2) | 133 (59.6) |
| 0.35 | Open | 157 (87.7) | 131 (58.7) |
| 0.35 | Close | 156 (87.2) | 131 (58.7) |
| 0.40 | Open | 157 (87.7) | 131 (58.7) |
| 0.40 | Close | 156 (87.2) | 130 (58.3) |

Percentages are based on 179 HC and 223 MCI trial rows per condition.

**B. Conditional kinetic and Group × Trial results**

| **R² threshold** | **Condition** | **Outcome** | **N trials** | **Group β (MCI vs HC)** | **P group** | **Group × Trial F** | **P interaction** | **Linear interaction β** | **P linear** |
| --- | --- | --- | --- | --- | --- | --- | --- | --- | --- |
| 0.30 | Open | LogTau | 276 | 0.079 | 0.407 | 0.406 | 0.667 | -0.090 | 0.399 |
| 0.30 | Open | t_onset_ | 276 | 0.112 | 0.003 | 8.611 | <0.001 | -0.162 | <0.001 |
| 0.30 | Close | LogTau | 280 | -0.072 | 0.388 | 0.058 | 0.943 | -0.020 | 0.806 |
| 0.30 | Close | t_onset_ | 280 | -0.028 | 0.678 | 8.909 | <0.001 | 0.233 | <0.001 |
| 0.35 | Open | LogTau | 275 | 0.055 | 0.557 | 0.292 | 0.747 | -0.062 | 0.557 |
| 0.35 | Open | t_onset_ | 275 | 0.107 | 0.005 | 8.138 | <0.001 | -0.158 | <0.001 |
| 0.35 | Close | LogTau | 278 | -0.050 | 0.553 | 0.160 | 0.853 | -0.029 | 0.721 |
| 0.35 | Close | t_onset_ | 278 | -0.024 | 0.722 | 8.222 | <0.001 | 0.222 | <0.001 |
| 0.40 | Open | LogTau | 275 | 0.055 | 0.557 | 0.292 | 0.747 | -0.062 | 0.557 |
| 0.40 | Open | t_onset_ | 275 | 0.107 | 0.005 | 8.138 | <0.001 | -0.158 | <0.001 |
| 0.40 | Close | LogTau | 277 | -0.059 | 0.485 | 0.199 | 0.820 | -0.043 | 0.593 |
| 0.40 | Close | t_onset_ | 277 | -0.022 | 0.746 | 8.545 | <0.001 | 0.228 | <0.001 |

Group β is the covariate-adjusted MCI-minus-HC coefficient: in seconds for t_onset_ and on the natural-log scale for LogTau. The categorical Group × Trial statistic treats Trial as a factor; the linear interaction coefficient uses Trial coded 0, 1 and 2. P group, P interaction and P linear are the unadjusted sensitivity-model P values shown for each R² threshold; prespecified Holm-adjusted primary comparisons are reported in main Tables 2 and 3 and Supplementary Table S3. The primary quality-control threshold is R² > 0.35.

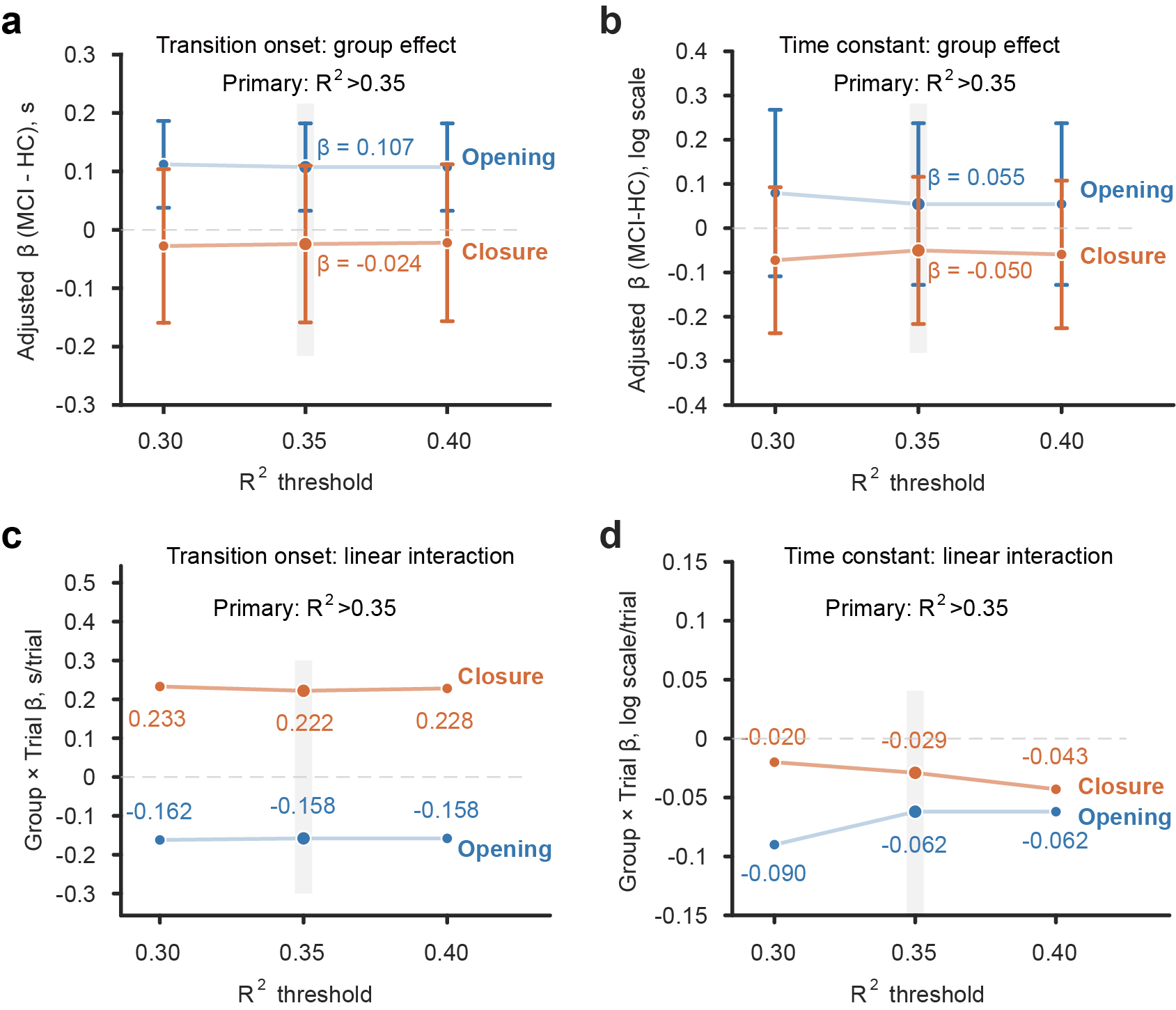

**Supplementary Figure S3 | Sensitivity of kinetic results to the curve-fitting quality threshold.**

Sensitivity of kinetic estimates to the FitQC threshold (R²>0.30, >0.35 or >0.40). **a,** Adjusted MCI-minus-HC effects on t_onset_; **b,** corresponding effects on LogTau. Vertical bars show 95% confidence intervals. **c,d,** Linear Group × Trial coefficients for t_onset_ (c) and LogTau (d). Blue denotes eye opening and orange eye closure; grey shading marks the primary threshold of R²>0.35. Values are reported in Supplementary Table S4.

### Supplementary Result 5. Associations between repetition-related EEG changes and cognitive performance

Within the MCI group, none of the six prespecified repetition-related EEG changes remained significantly associated with MoCA after Holm correction (Supplementary Table S5 and Supplementary Figure S4). In addition to the prespecified ΔLogTau analysis, we examined the untransformed change in time constant (Δτ=τ_T3_−τ_T1_).

During eye opening, Δτ showed a non-significant negative association with MoCA (Spearman ρ=-0.303; Holm-adjusted P=0.154). The association remained non-significant after adjustment for age, sex and years of education (β=-1.80, 95% CI -4.91 to 1.32; Holm-adjusted P=0.533). During eye closure, Δτ was not associated with MoCA (ρ=0.054; Holm-adjusted P=0.757), and the adjusted regression was also non-significant. These findings were consistent with the primary analyses based on ΔLogTau.

**Supplementary Table S5 | Associations between repetition-related EEG changes and MoCA scores within the MCI group**.

**A. Prespecified delta measures**

| **Parameter** | **N** | **Spearman ρ** | **P** | **Holm P** | **Adjusted β** | **95% CI** | **Holm P (regression)** |
| --- | --- | --- | --- | --- | --- | --- | --- |
| Open ΔLogER | 70 | 0.032 | 0.792 | 1.000 | -0.002 | -0.663, 0.659 | 1.000 |
| Close ΔLogER | 70 | 0.244 | 0.042 | 0.250 | 0.611 | -0.059, 1.281 | 0.472 |
| Open ΔLogTau | 35 | -0.297 | 0.084 | 0.418 | -0.621 | -1.596, 0.353 | 1.000 |
| Close ΔLogTau | 35 | 0.000 | 1.000 | 1.000 | -0.204 | -1.610, 1.203 | 1.000 |
| Open Δt_onset_ | 35 | 0.288 | 0.094 | 0.418 | 1.598 | -0.819, 4.016 | 1.000 |
| Close Δt_onset_ | 35 | -0.174 | 0.316 | 0.949 | -1.080 | -3.218, 1.057 | 1.000 |

Changes were defined as T3−T1. Adjusted regression models included age, sex and years of education. Holm correction was applied across the six prespecified association tests. MoCA, Montreal Cognitive Assessment; MCI, mild cognitive impairment.

**B. Δτ sensitivity analysis**

| **Parameter** | **N** | **Spearman ρ** | **Holm P** | **Adjusted β** | **95% CI** | **Holm P (regression)** |
| --- | --- | --- | --- | --- | --- | --- |
| Open Δτ | 35 | -0.303 | 0.154 | -1.799 | -4.912, 1.315 | 0.533 |
| Close Δτ | 35 | 0.054 | 0.757 | 0.245 | -3.605, 4.095 | 0.902 |

Adjusted regression models included age, sex and years of education. Δτ=τ_T3_−τ_T1_ is the untransformed T3−T1 change in the fitted time constant.

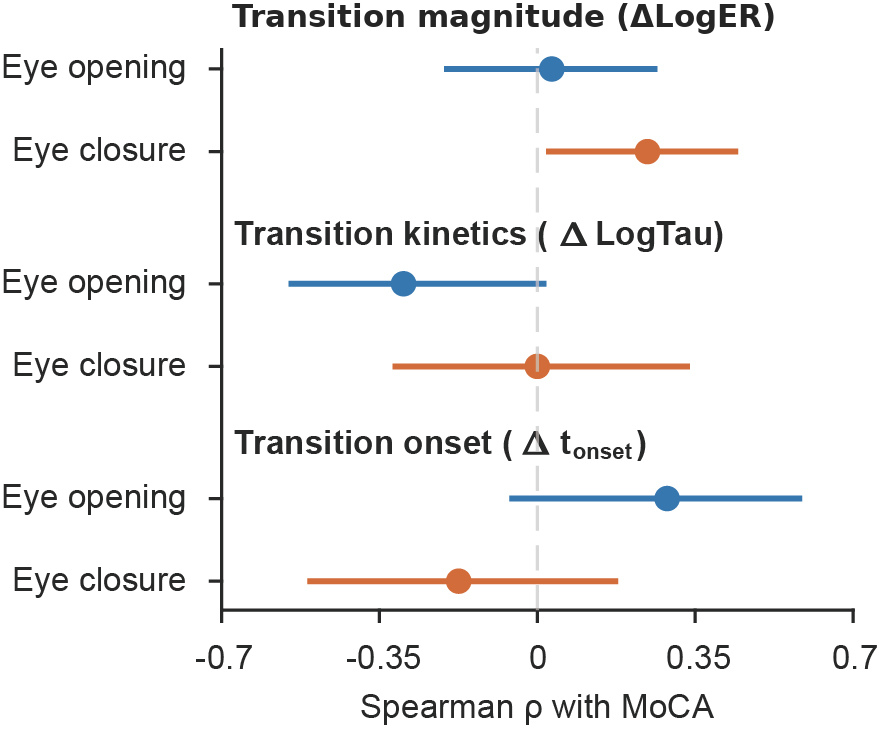

**Supplementary Figure S4 | Associations between repetition-related changes in alpha state-transition measures and MoCA scores in participants with MCI**.

Spearman associations between MoCA and T3−T1 changes in alpha-transition measures within MCI. Points show Spearman's ρ and horizontal bars participant-level bootstrap 95% confidence intervals (5,000 resamples) for ΔLogER, ΔLogTau and Δt_onset_ during eye opening (blue) and closure (orange). The dashed line denotes ρ=0. Sample sizes were n=70 for ΔLogER and n=35 for each kinetic outcome; Holm-adjusted results are reported in Supplementary Table S5.
